# Impact of Aortic Valve Leaflets Calcium Volume and Distribution on Post-TAVR Conduction Abnormalities

**DOI:** 10.1101/2025.05.04.25326961

**Authors:** Symon Reza, Brandon Kovarovic, Danny Bluestein

**Affiliations:** Department of Biomedical Engineering, Stony Brook University, Stony Brook, New York, USA

**Keywords:** TAVR, FEA, Conduction abnormality, PPI, Leaflet calcification, Living Heart Model

## Abstract

**Introduction:** Transcatheter aortic valve replacement (TAVR) is increasingly used to treat aortic stenosis, including in low-risk patients. However, post-procedural cardiac conduction abnormalities (CCA), often requiring permanent pacemaker implantation (PPI), remain a concern. This study investigates how the volume and distribution of aortic leaflet calcium deposits influence the risk of post-TAVR CCA.

**Methods:** An electromechanically coupled four-chamber beating heart model was used to simulate TAVR with a self-expandable Evolut® 26 mm device. Five virtual patient scenarios were modeled with varying calcium volumes and distributions: no calcium, uniform distribution (3 Calc), and isolated calcification on the left coronary leaflet (LCL), right coronary leaflet (RCL), or non-coronary leaflet (NCL). Electrical conduction was simulated using a monodomain model and coupled with structural mechanics to evaluate tissue-device interactions. Metrics included principal stress, contact pressure, and contact pressure index (CPI) over three cardiac cycles.

**Results:** Larger calcium volumes and specific leaflet distributions increased stress and contact pressure near the atrioventricular node. The LCL model exhibited the highest mechanical stress and peak contact pressure (13.1 kPa), while the NCL model showed the lowest (6.42 kPa). The RCL model had intermediate values. Elevated contact pressure and stress in the LCL case suggest an increased risk of conduction disruption and PPI.

**Conclusion:** Leaflet calcium deposit volume and distribution significantly influence mechanical stress and contact dynamics near the conduction system following TAVR. These insights support the integration of clinical data, such as leaflet calcium volume and distribution into pre-procedural planning to personalize risk assessment and improve patient outcomes.

**Highlights:**

- Beating heart model used to assess post-TAVR conduction abnormality risk.
- Higher calcium volume increases contact pressure near conduction pathways.
- Left coronary leaflet calcification linked to elevated post-TAVR CCA risk.
- Dynamic simulations show stress fluctuations during cardiac cycles.
- Findings support personalized TAVR planning to reduce PPI need.

## 1. Introduction

Aortic stenosis has traditionally been treated with surgical aortic valve replacement (SAVR), while transcatheter aortic valve replacement (TAVR) has emerged as a minimally invasive alternative. With comparable hemodynamic outcomes, TAVR is now widely adopted in high- and intermediate-risk patients and has recently been approved for low-risk patients. However, this expansion raises concerns about complications such as paravalvular leakage, leaflet thrombosis, valve degeneration, and notably, cardiac conduction abnormalities (CCAs). Among these, CCAs are particularly persistent and often require permanent pacemaker implantation (PPI) [1–3], limiting TAVR’s broader application. CCAs typically result from mechanical injury to the atrioventricular conduction system near the interleaflet triangle between the non-coronary and right coronary leaflets. PPI has been linked to increased mortality and heart failure hospitalizations [2, 4], making CCA risk prediction critical for improved pre-procedural planning.

Several clinical studies have identified risk factors for post-TAVR CCA, including preexisting right bundle branch block (RBBB), deeper implantation, short membranous septum length, low ΔMSID, and calcium burden. However, findings—especially on the volume and distribution of leaflet calcification—have been inconsistent. Some link high calcium on the left coronary leaflet (LCL) with increased PPI rates [5, 6], while others implicate the non-coronary (NCL) [7] or right coronary leaflets (RCL) [8]. These discrepancies complicate clinical decision-making and highlight the need for more precise predictive tools. Moreover, anatomical variability in the device landing zone limits the reliability of purely anatomical and procedural predictors. Since post-TAVR CCA depends on localized, dynamic interactions between the prosthesis and conduction fibers, computational modeling offers a promising way to enhance predictive accuracy.

Computational tools have been widely used to study TAVR complications like paravalvular leakage, thrombosis, and valve degeneration [9–14]. Patient-specific models have also been applied to analyze post-TAVR CCA risk, showing that elevated contact pressure or strain on the His bundle correlates with conduction damage [15–17]. Recently, we introduced a beating heart electro-mechanical model to simulate post-TAVR CCA risk under dynamic conditions [18]. While more physiologically accurate, this method is computationally demanding and not yet scalable to patient-specific workflows. However, it holds value in helping clinicians interpret how measurable parameters—such as calcium volume and distribution—affect post-TAVR outcomes and may reconcile inconsistent findings across clinical studies.

The Living Human Heart Model (a, b) is a high-fidelity, electromechanically coupled, four-chamber adult male heart developed under the SIMULIA Living Heart Project. It has been used to study various cardiovascular conditions and interventions, including TAVR, mitral repair, and LVADs [18–22]. Prior studies have mapped calcium patterns on native leaflets [23] and Halevi et al. [24] used the reverse calcification technique (RCT) to identify common geometries, showing deposits mainly along the coaptation line (Figure 2), consistent with Thubrikar et al. [23].

In this study, we used the Living Human Heart Model to simulate common calcium distribution patterns by structurally modifying the native valve to introduce synthetic leaflet calcifications with varying volumes and distributions (Figure 1c).Our goal was to evaluate their role in increasing CCA risk and provide insights that support clinically relevant, data-driven decision-making using readily available parameters such as calcium burden.

**Figure 1:**
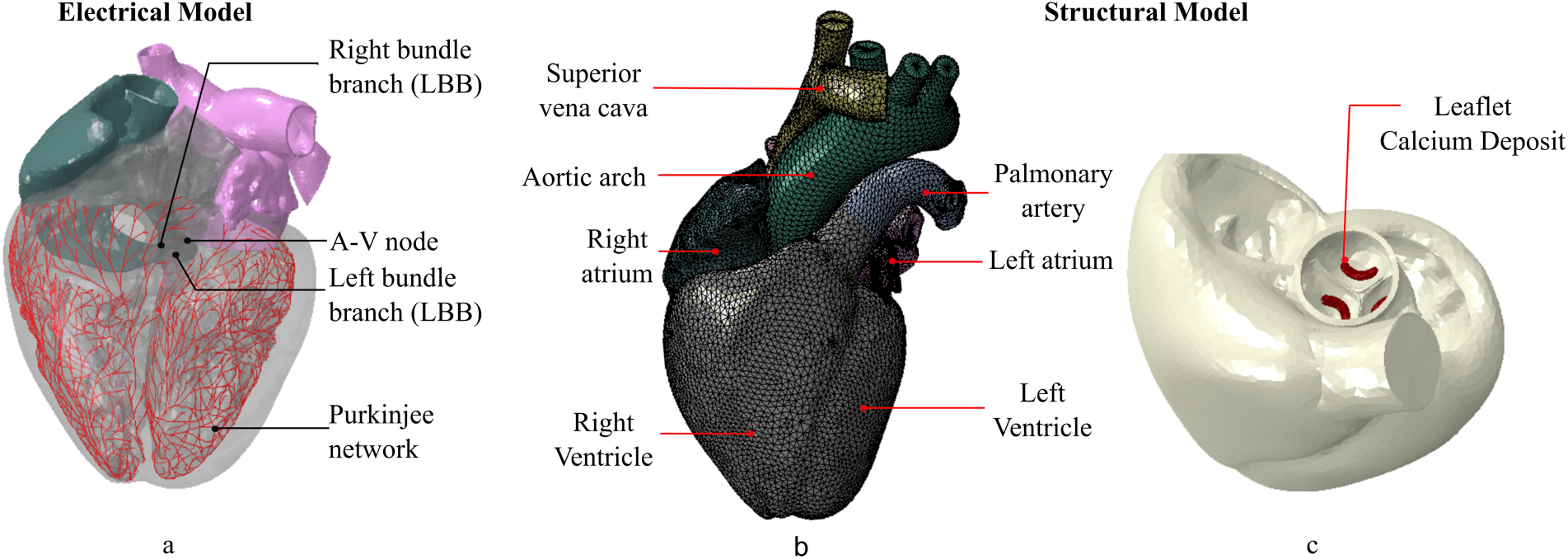
Illustration of the (a) electrical model, (b) finite element model, of the 4-chamber full heart model (Adapted with permission from Reza et al. Biomechanics and Modeling in Mechanobiology [18]), and (c) diseased heart with leaflet calcification.

**Figure 2:**
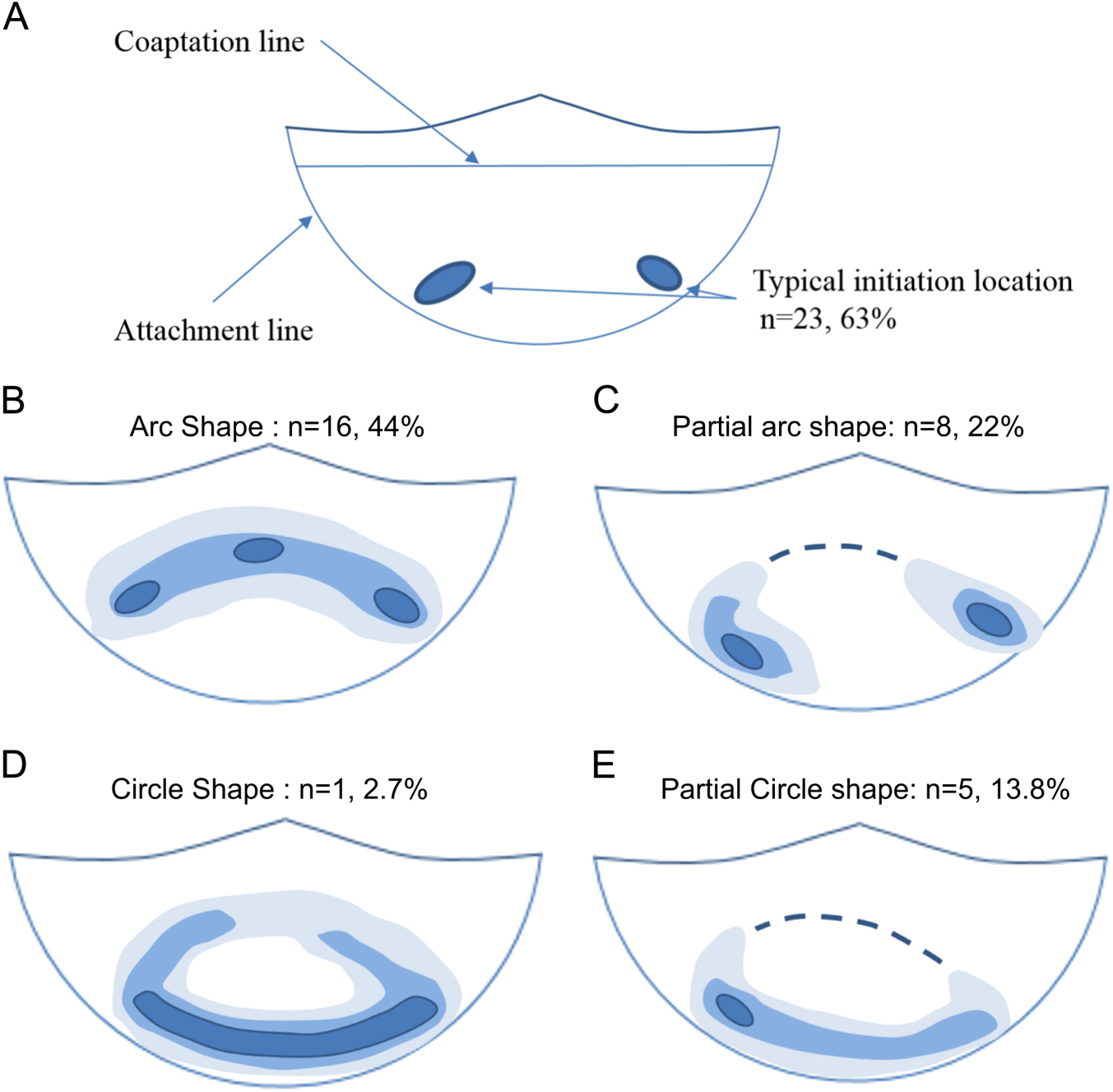
Common patterns of calcium deposits (Reproduced with permission from Halevi et al., Journal of Biomechanics [24])

## 2. Methods

A dynamic, high-fidelity four-chamber human heart model developed under the SIMULIA Living Heart Model [25], was used as the foundation for this study. This model employs a sequentially coupled electro-mechanical approach to simulate realistic cardiac function. Electrophysiological simulations of healthy cardiac conduction were performed first, and the resulting activation patterns were coupled with the structural model to replicate mechanical heart behavior. An Evolut® 26 mm TAVR device (Medtronic, Inc., Minneapolis, MN) was deployed within this beating heart model under five different aortic leaflet calcium deposit conditions. A 6 mm implantation depth—within the clinically recommended 3–6 mm range—was selected as the nominal case. The membranous septum (MS) length in the model was 5.5 mm, yielding ΔMSID values of −1.5 mm (aortic), +0.5 mm (nominal), and +2.5 mm (ventricular).

### 2.1. Calcium deposit model and distribution

To investigate how calcium volume and distribution affect post-TAVR CCA risk, we created parametric calcium deposit models based on commonly observed leaflet calcification shapes. Each leaflet-specific model was fitted to the Living Heart Model geometry with a average calcium volume of 85 mm³. Five virtual patient scenarios were created (Figure 3). : (1) a healthy case with no calcium deposits, (2) a case with uniform deposits across all three leaflets (3 Calc), and three cases with localized calcium on the non-coronary (NCL), right coronary (RCL), or left coronary (LCL) leaflet, respectively.

**Figure 3:**
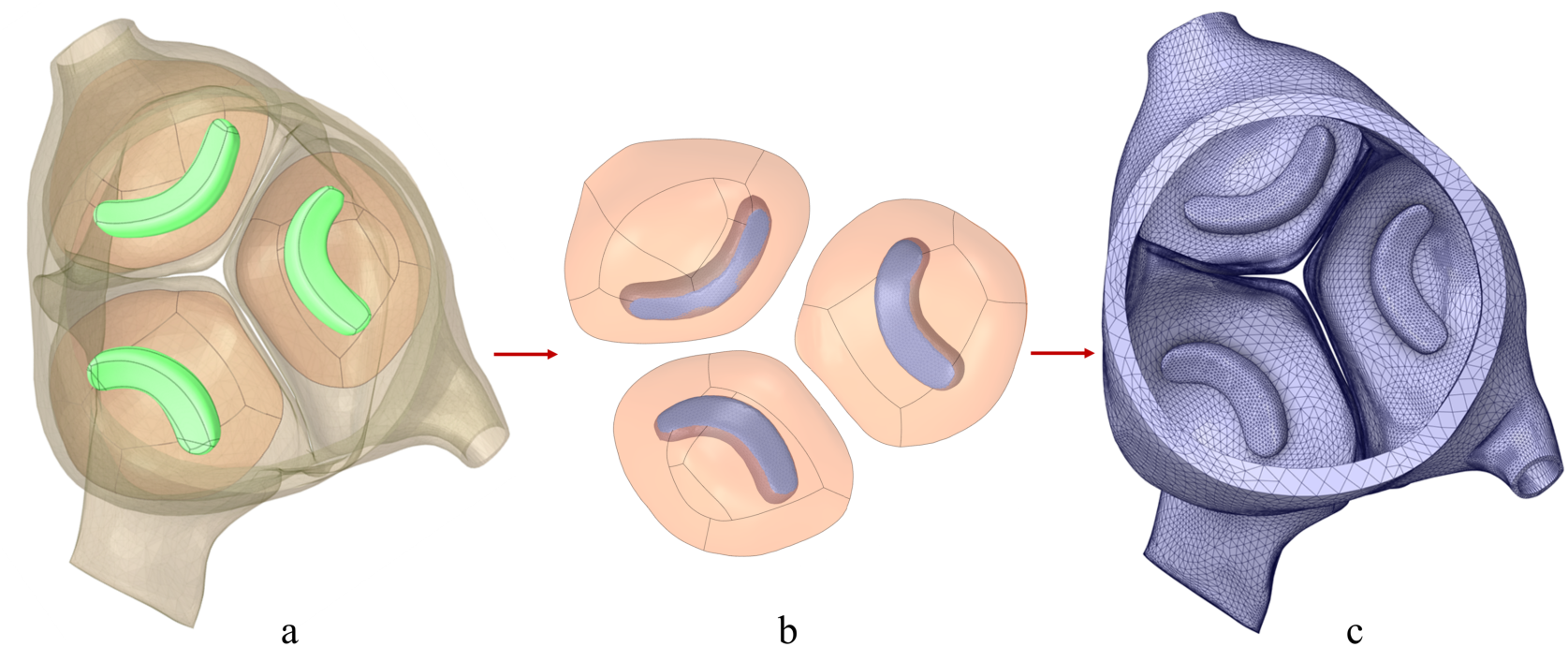
Idealized leaflet calcium deposit model creation, (a) CAD model creation, (b) leaflet surface fitting of the CAD models of the leaflet calcium deposits, and (c) meshed calcium deposit models.

### 2.2. Modeling cardiac electrophysiology

Cardiac conduction was simulated in Abaqus/Standard using a monodomain reaction-diffusion model. Electrical signals propagated through both myocardial tissue and a 1D Purkinje fiber network, based on geometries from literature [25]. The model accounted for isotropic and anisotropic conduction and was solved using a staggered implicit Euler scheme over 500 ms of cardiac activity, beginning at 70% ventricular diastole. The heart was discretized into 297,635 tetrahedral elements, while the Purkinje network consisted of 10,757 1D elements, coupled to the myocardium through resistor-like conduction links. Electrical activation results were mapped to the mechanical model as boundary conditions, ensuring realistic electromechanical behavior. The implementation followed methods in the literature [26, 27].

### 2.3. Structural modeling

The structural model included detailed representations of both active and passive myocardial responses. Passive behavior followed an anisotropic hyperelastic formulation [29], with stiffness calibrated to match experimental strain data [26]. Active contraction was modeled using a time-varying elastance framework [27], modulated by a contractility factor to replicate physiological ventricular twist and stroke volume. Fiber orientations ranged from −60° (epicardium) to +60° (endocardium) [28], capturing the heart’s layered structure. Model calibration ensured accurate ventricular deformation, chamber pressure-volume relationships, and realistic wall motion [29]. Further implementation details are available in the SIMULIA Living Heart Human Model User Guide (2018) [30].

### 2.4. Simulation parameters

The simulation was conducted in two stages: electrical and structural. In the first stage, cardiac conduction was simulated as described in Section 2.1. The resulting electrical activation data were applied as boundary conditions in the structural simulation to replicate realistic cardiac contraction using Abaqus Explicit 2019 (SIMULIA, Dassault Systèmes, Providence, RI). The structural phase included six sequential steps: crimping, placement, deployment, preload, beat, and recovery, spanning three cardiac cycles (Figure 4).

**Figure 4:**
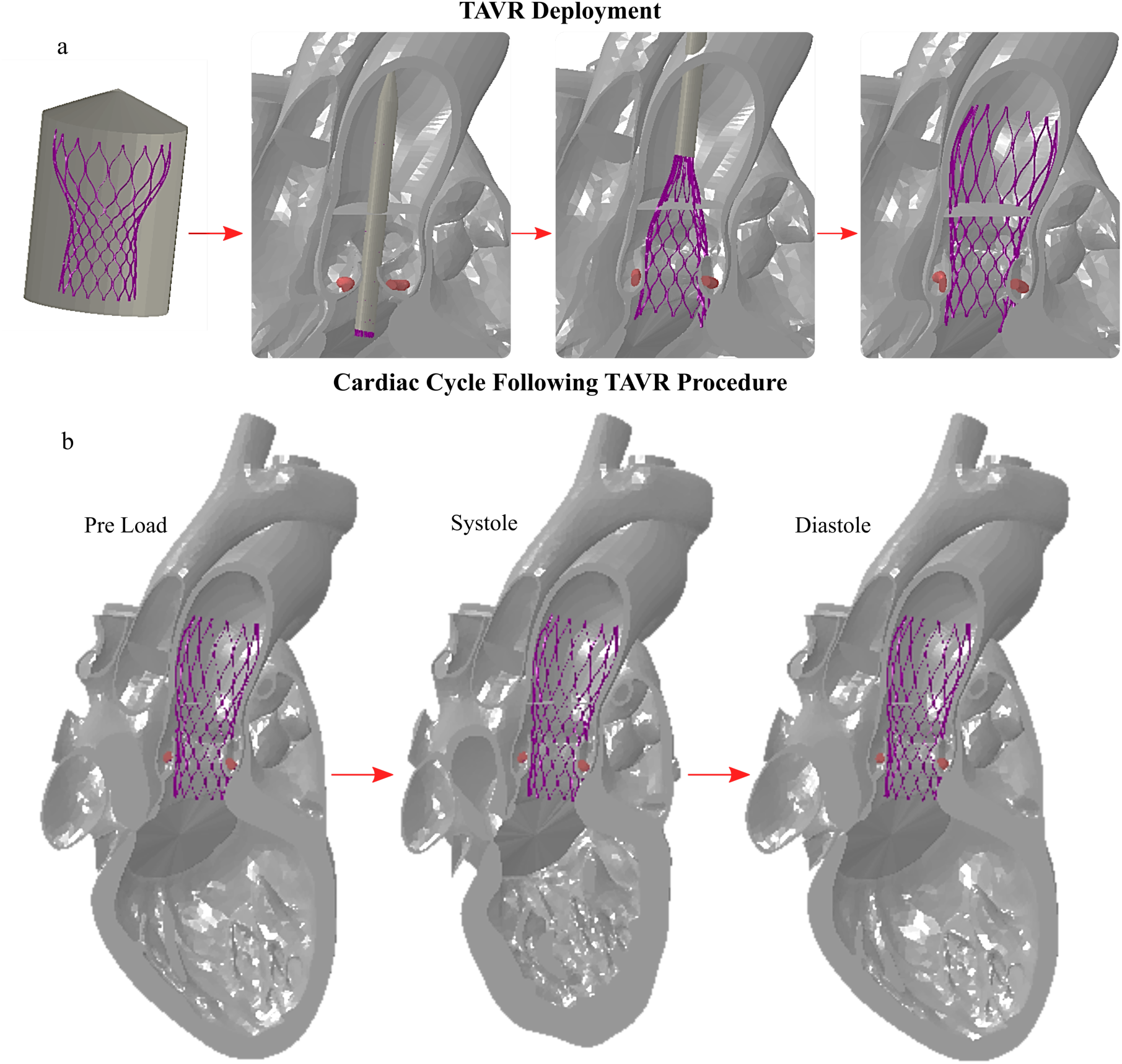
(a) TAVR implantation sequence from crimping to deployment, and (b) cardiac cycles following the procedure.

The TAVR stent was modeled using a structured hexahedral mesh and assigned a superelastic Nitinol material model [32, 33], with further material details available in [17]. The His bundle (HB) is anatomically located between the atrioventricular membranous septum (MS) and the posterior crest of the muscular septum, beneath the interleaflet triangle between the non-coronary (NCL) and left-coronary (LCL) leaflets [34]. Accordingly, the area of interest (AOI) for stress and contact analysis was defined just below the MS, in the region between the NCL and LCL (Figure 5), following the methodology outlined in Rocatello et al. [15].

**Figure 5:**
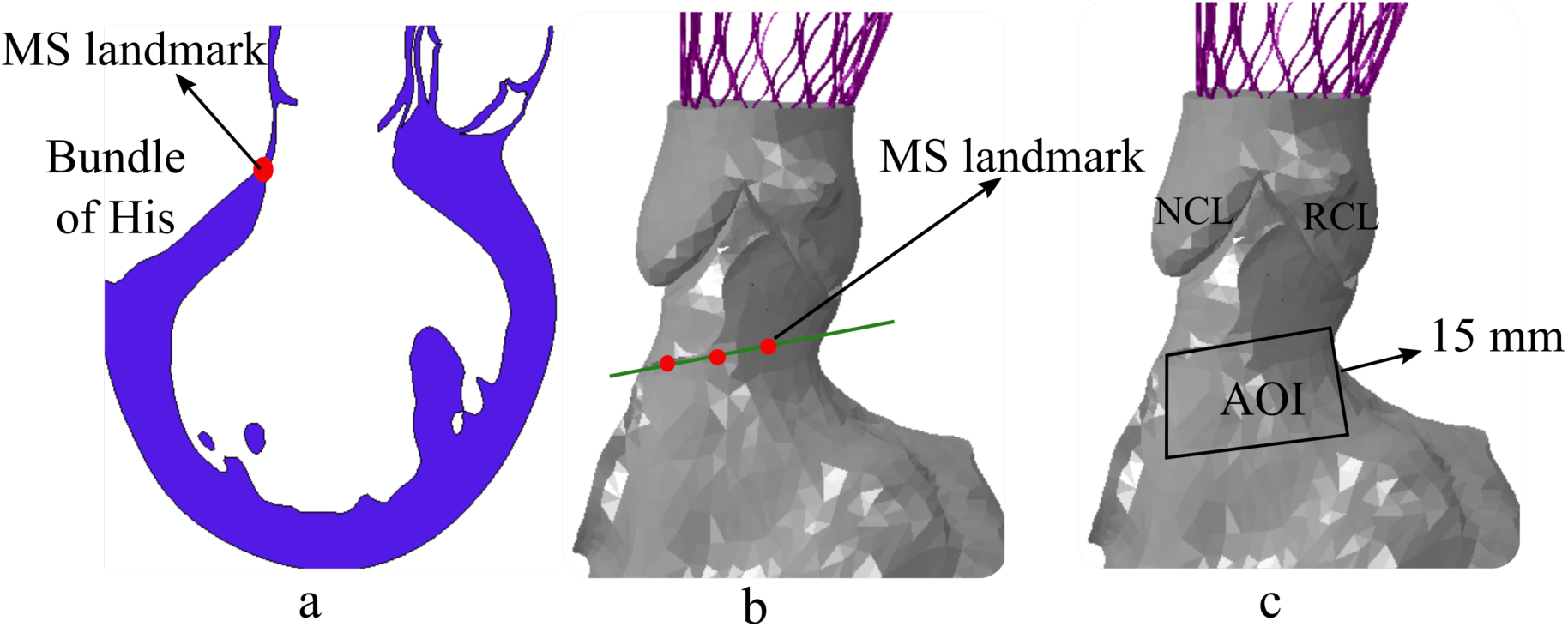
Illustration of the MS landmark from (a)) long axis view. Identified MS landmark in shown in green (b) and identified area of interest is confined to a black box (c).

## 3. Results

The supra-annular Evolut® 26 mm device was implanted inside the aortic annulus of the modified Living Heart Model with five varying aortic leaflet calcium deposit volume and distribution (Figure 6). To assess prosthesis–tissue interaction, three cardiac cycles were simulated following deployment. Hoop stresses on the area of interest (AOI) were evaluated for each case and compared to each other. Additionally, contact force, contact pressure, and the contact pressure index (CPI) were analyzed across all virtual patient scenarios.

**Figure 6:**
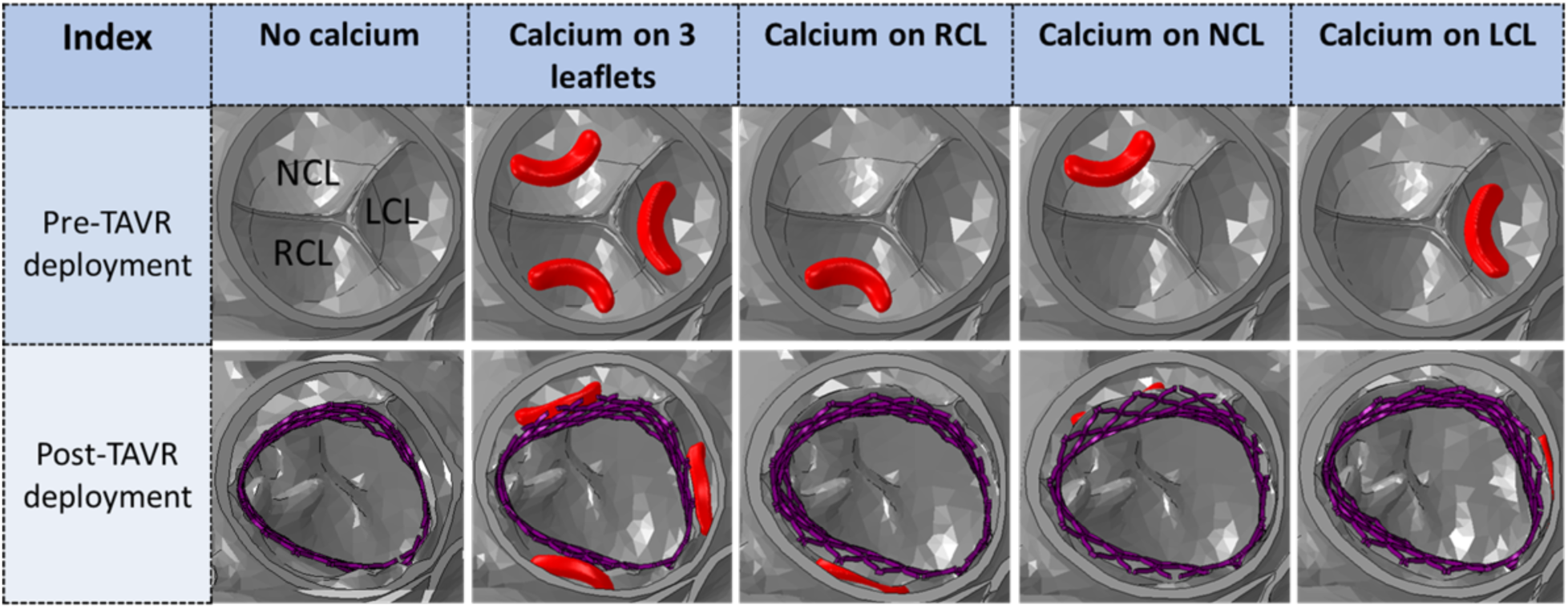
Pre-TAVR patient cases of various leaflet calcium deposit volume and distribution

### 3.1. Impact of calcium deposit volume

To evaluate the impact of calcium deposit volume on post-TAVR biomechanics, von Mises stress, principal stress components, and contact-based parameters—such as contact pressure and pressure index—on the area of interest (AOI) were analyzed. Following TAVR implantation, all three principal stress components increased. Over three cardiac cycles, the 3 Calc model showed stress increases of 20.95% (axial), 5.49% (radial), and 24.13% (hoop), while the No Calc model exhibited smaller increases of 16.52%, 7.19%, and 21.49%, respectively, compared to the healthy case.

Contact-based metrics also revealed substantial differences. During device deployment, maximum contact pressure peaked and stabilized by preload but fluctuated with each heartbeat post-deployment—reaching a peak from late systole to early diastole. The 3 Calc model experienced the highest contact pressure, peaking at 10.3 kPa with a mean of 1.93 kPa, while the No Calc model recorded a lower peak of 5.69 kPa and a mean of 0.52 kPa (Figure 7a).

**Figure 7:**
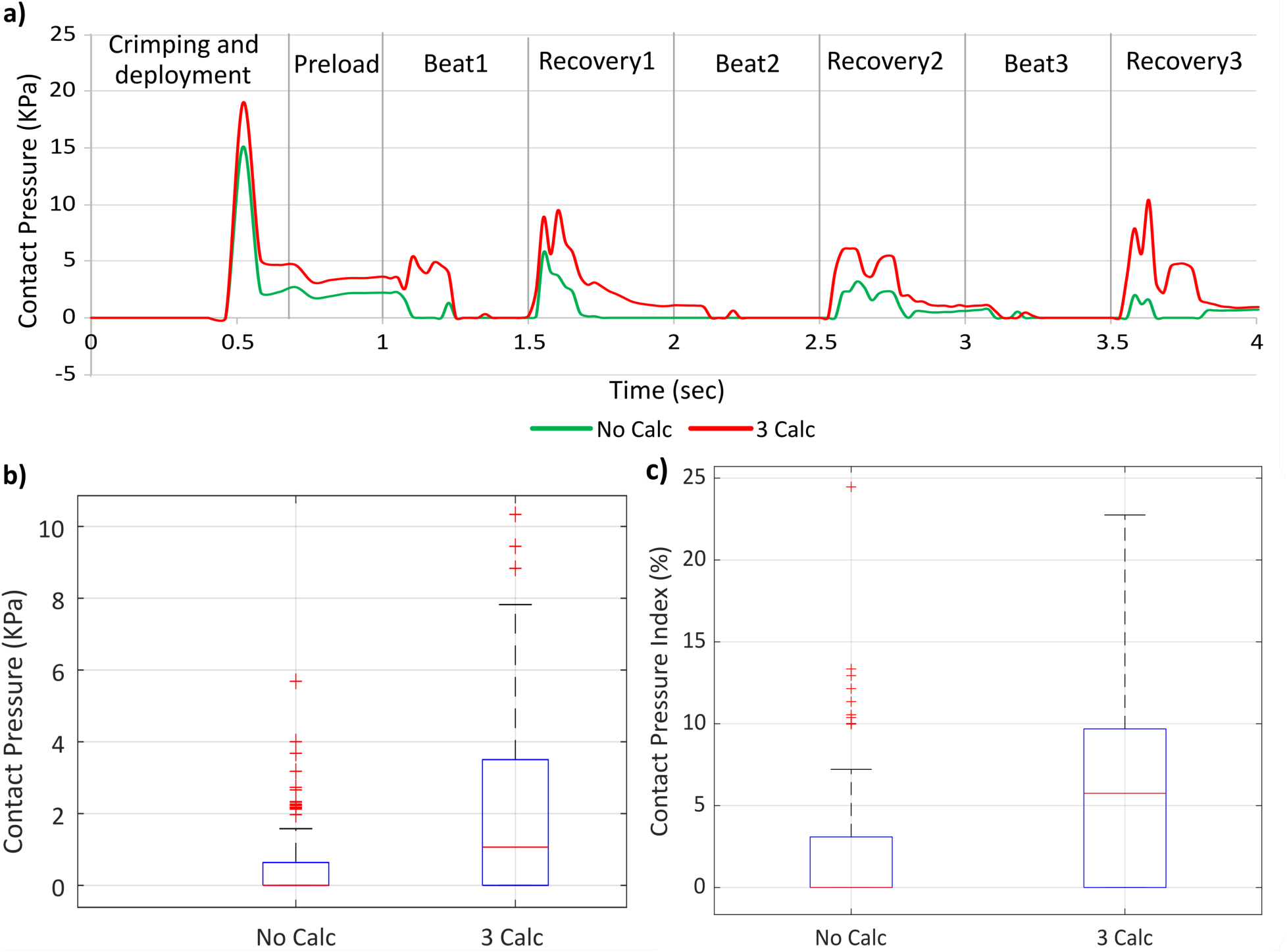
Effects of calcium deposit volume, (a) Contact pressure on the AOI at each step;(b) Box plots of contact pressure at the AOI across three cardiac cycles following TAVR implantation for two different calcium deposit volume, and (c) Contact Pressure Index (CPI) during the three cardiac cycles.. A Mann-Whitney U test comparing contact pressures across calcium deposit volume yielded a statistically significant difference (P < 0.001).

The cumulative contact pressure over three cardiac cycles was 234.61 kPa for the 3 Calc model and 65.88 kPa for the No Calc model (Figure 7b). Additionally, contact area analysis revealed a slightly higher contact pressure index for the 3 Calc model (26.33%) compared to the No Calc model (25.2%) (Figure 7c), highlighting how increased calcium volume elevates mechanical stress and contact forces on the conduction system region.

### 3.2. Impact of calcium deposit distribution

A similar trend was observed with varying calcium deposit distributions. A pronounced peak in maximum contact pressure occurred during the TAVR deployment phase, stabilizing by the end of deployment and preload. During subsequent heartbeats, contact pressure fluctuated cyclically, with peaks occurring from the late systole to early diastole in each cardiac cycle. The range of maximum principal stress components over three cardiac cycles was compared. The LCL Calc model showed increases of 15.96%, 8.38%, and 23% in axial, radial, and hoop stress, respectively. The RCL Calc model exhibited increases of 16.17%, 7.57%, and 22.64%, while the NCL Calc model showed smaller increases of 14.61%, 7.43%, and 19.42% in the same stress components.

Contact-based parameters also varied significantly. The LCL Calc model exhibited the highest contact pressure throughout the simulation, with a maximum of 13.1 kPa and a mean of 1.91 kPa. The RCL Calc model recorded a maximum of 9.41 kPa and a mean of 0.90 kPa, while the NCL Calc model had the lowest, with a peak of 6.42 kPa and a mean of 0.59 kPa (Figure 8a). The temporal sum of contact pressure across three cardiac cycles further reflected these differences, with values of 232.40 kPa for LCL, 110.72 kPa for RCL, and 73.24 kPa for NCL (Figure 8b). Contact area analysis showed contact pressure indices of 25.67%, 20%, and 25.80% for the LCL, RCL, and NCL Calc models, respectively (Figure 8c), highlighting how both calcium distribution and leaflet location influence prosthesis–tissue interaction.

**Figure 8:**
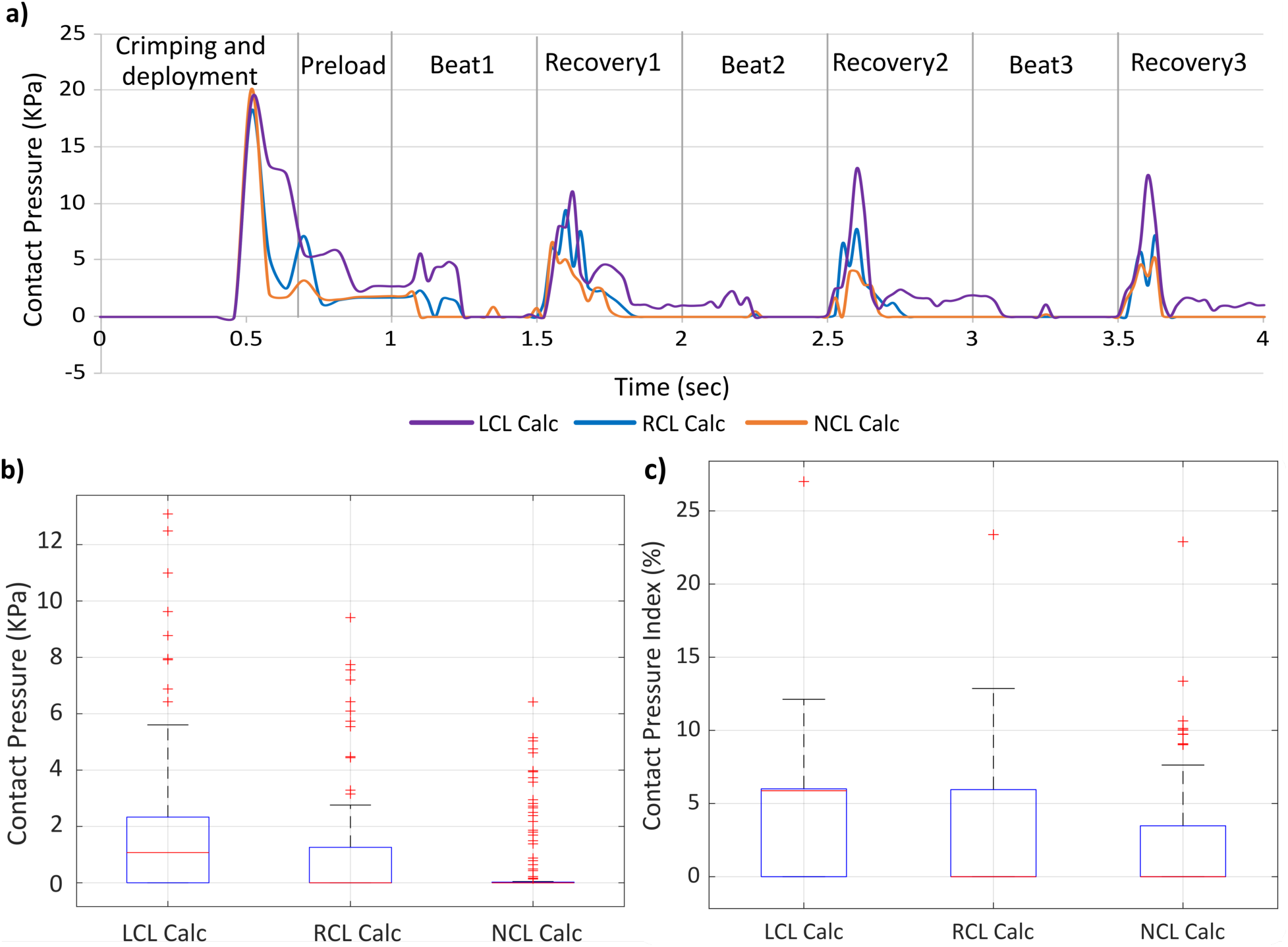
Effects of calcium deposit distribution, (a) Contact pressure on the AOI at each step, (b) Box plot diagrams of contact pressure at the AOI during three cardiac cycles following TAVR implantation for three distributions of calcium deposits, and (c) CPI during three cardiac cycles. Kruskal-Wallis test was performed between the contact pressures and contact pressure indexes three distributions of calcium deposits yielding P<0.001.

## 4. Discussion

Post-TAVR cardiac conduction abnormalities (CCA) remain a critical complication, often leading to permanent pacemaker implantation (PPI). PPI is associated with increased mortality and heart failure hospitalizations, impacting the broader success of TAVR. This complication underscores the critical need of identifying and understanding the anatomical and procedural factors contributing to post-TAVR CCA. Key contributors to these conduction abnormalities include prosthesis implantation depth, membranous septum (MS) length, (ΔMSID), preexisting RBBB, and calcium burden. Understanding the complex interplay between post-procedural CCA, preexisting conduction abnormalities, and the interaction between the TAVR prosthesis and cardiac conduction fibers remain a challenge. Previously, we introduced an innovative approach using a high-fidelity, electro-mechanically coupled beating heart model, which provided deeper insights into the relationship between these abnormalities and factors such as implantation depth and preexisting cardiac asynchronies [18]. However, that study was limited to parametric modeling. Furthermore, clinical studies sought to establish a link between commonly accessible clinical data, such as the volume and distribution of calcium deposits on the aortic leaflets, and the new onset of post-TAVR CCA, but the findings were inconclusive, prompting us to numerically investigate how the volume and distribution of aortic leaflet calcium deposits influence the onset of CCA. Accordingly, in this study, we parametrically modified the model to evaluate the relative risk of post-TAVR CCA based on the volume and distribution of leaflet calcium deposits.

The results of this study show that both the volume and distribution of aortic leaflet calcium deposits significantly influence the risk of post-TAVR CCA. When calcium is concentrated on certain leaflets, it alters prosthesis expansion, increasing contact pressure and mechanical stress within the area of interest (AOI) throughout the cardiac cycle. This observation is consistent with prior research suggesting that high volume of calcification may contribute to mechanical damage to the conduction system and increase the relative risk of post TAVR CCA [8, 31–33]. Specifically, LCL calcification led to the highest stress and pressure within the AOI, as the device was deflected toward the RCL–NCL junction. This suggests that calcification on the LCL may heighten the risk of post-TAVR CCA, particularly for self-expandable TAVR procedures. Conversely, calcium deposits on the RCL led to lower stress and contact pressure than in the LCL Calc model yet remained significantly higher than those in the NCL Calc model.

Additionally, the curvature of the aortic arch pushes the top crown of the Evolut® device toward the LCL, causing the calcified leaflets to act as a pivotal plane. This leads to rotational forces twisting the bottom crown, shifting it towards the opposite direction-approximately toward the AOI, while larger deposits on the NCL slightly shift the prosthesis toward the RCL, shielding the AOI from contact with the prosthesis. It is important to note that the study used a parametric model for calcium deposits, whereas in reality, the shape of calcium deposits tends to be uneven. This suggests that similar calcium distributions with varying shapes could yield different outcomes. Therefore, patient-specific modeling may further increase the accuracy for assessing the risk of post-TAVR CCA.

Previous clinical studies have investigated leaflet calcium distribution as a predictor of post-TAVR CCA and the need for PPI. Many of these studies focused on either balloon-expandable devices or both balloon- and self-expandable devices. For example, Mauri et al. [34] and Bianchini et al. [8] examined balloon-expandable devices and found that a higher calcium volume at the left ventricular outflow tract (LVOT) below the LCL and RCL, respectively, was associated with an increased need for PPI. Fujita et al. [5] studied both balloon- and self-expandable devices and identified high calcium distribution on LCL as a contributing factor to the need for PPI. In contrast, Pollari et. al. [35] and Gama et. al. [36] found calcium deposits below the NCL at the LVOT to be linked with the need for PPI. Veulemans et al. [6] and Sharma et. al. [7] focused on self-expandable TAVR systems and identified high calcification on the LCL as a dependent variable. The current study observed that higher calcium volumes in the LCL increase the risk of post-TAVR CCA which aligned with the observation from Veulemans et al. [6] and Sharma et. al. [7] both of the studies focusing on self-expandable devices. These findings suggest that mixing balloon-expandable and self-expandable devices in studies could yield conflicting results, and leaflet calcification may not be a uniform risk factor across device types, as the material properties, shapes, and implantation techniques of these devices differ significantly. Furthermore, well-established anatomical and procedural factors such as implantation depth, membranous septum length (ΔMSID), and preexisting RBBB are likely to play a central role in post-TAVR outcomes, while leaflet calcium volume and distribution may act as secondary risk enhancers rather than primary drivers.

In summary, this study aimed to clarify the conflicting clinical findings on the relationship between specific calcification patterns and the need for post-procedural pacemaker implantation (PPI). By closely examining the interaction between the TAVR device and the AOI across varying calcium deposit volumes and distributions—while keeping other anatomical and procedural parameters constant, the study provides valuable insights into this complex association. The results indicated that the volume of calcium deposits on the aortic leaflets may increase the risk of post TAVR CCA in self-expandable TAVR devices. High calcium deposit volume on the LCL may increase the risk compared to RCL and NCL for self-expandable TAVR devices. Calcium deposits on the LVOT and balloon expandable devices need to be studied separately to understand the device type specific effect of calcium deposits on the onset of CCA and the need for PPI.

The dynamic fluctuations in contact pressure throughout the cardiac cycle emphasize the need to account not only for static anatomical features but also for the complex dynamic biomechanical interactions between the TAVR prosthesis and surrounding cardiac structures during the entire cardiac cycle. Particularly, variations in contact pressure during the transition from late systole to early diastole may indicate transient mechanical stresses that increase the likelihood of the new onset of CCA following TAVR procedures. While traditional clinical parameters are essential, the integration of anatomical features and high-fidelity simulations allows for a more nuanced understanding of the biomechanical interactions that underlie post-TAVR complications. This approach could lead to more personalized pre-procedural planning, potentially reducing the incidence of CCA and improving overall patient outcomes. Overall, the findings suggest that clinicians should consider both volume and spatial pattern of calcium, especially on the LCL, when using Nitinol-based, supra-annular TAVR devices. These insights may improve risk stratification and guide personalized TAVR strategies.

## 5. Limitations

This study used idealized calcium shapes and average volumes to assess post-TAVR CCA risk. While this enabled controlled comparisons, incorporating patient-specific calcium geometries would improve clinical applicability. Simulations employed a unidirectional electro-mechanical coupling using the Living Human Heart Model (LHHM), excluding mechano-electrical feedback. Prior research indicates this omission has a minimal impact on conduction velocity [37] supporting the model’s validity for this application. The use of a standard healthy male heart model provides consistency but may not reflect anatomical variability in typical TAVR patients. Adapting simulations to patient-specific heart models will allow more accurate identification of mechanical thresholds linked to CCA risk. We plan to expand our current simulations to incorporate patient-specific models.

## 6. Conclusions

This study investigated how the volume and distribution of leaflet calcifications influence the risk of developing post-TAVR cardiac conduction abnormalities (CCA) that may necessitate a permanent pacemaker implantation (PPI). By evaluating the interaction between the TAVR device and the area of interest (AOI) under varying calcium patterns—while controlling for other anatomical and procedural variables, the results suggest that increased leaflet calcium burden, particularly on the left coronary leaflet (LCL), elevates CCA risk in self-expandable devices.

This study used of a beating heart model that captures the dynamic interplay between the device, tissue, and calcifications, offering a more physiologically realistic assessment compared to static device implantation analysis. This method supports personalized pre-procedural planning and can guide the design of next-generation TAVR devices aimed at reducing CCA risk and improving patient outcomes.

## Data Availability

The data is available within the body of the manuscript.

## Acknowledgment

The Dassault Systèmes SIMULIA Living Heart Project has provided the base framework of the 4-chamber beating heart model employed in this study as part of our academic collaboration with the company. The authors thank Dr. Matteo Bianchi for his contributions to the early stages of the project and for helping to develop the initial framework for analyzing CCAs in the beating heart, and Dr. Jiang Yao (Dassault Systèmes) for providing scientific insights.

SIMULIA Living Heart Project has provided the base framework of 4-chamber beating heart model employed in this study. The authors thank Dr. Matteo Bianchi for initiating the project and providing scientific insights.

## Acronyms

TAVR: Transcatheter aortic valve replacement
CCA: Cardiac conduction abnormality
MS: Membranous septum
PPI: Permanent pacemaker implantation
A-VI: Atrioventricular
AOI: Area of interest
HB: His bundle
CPI: Contact pressure index
CTA: Computed tomography angiography
LBBB: Left bundle branch block
RBBB: Right bundle branch block
RCL: Right coronary leaflet
NCL: Non-coronary leaflet
LCL: Left coronary leaflet
ΔMSID: Subtraction of implantation depth from the membranous septum length

